# Evidence of a Sjögren’s disease-like phenotype following COVID-19

**DOI:** 10.1101/2022.10.20.22281265

**Authors:** Yiran Shen, Alexandria Voigt, Laura Goranova, Mehdi Abed, David E. Kleiner, Jose O. Maldonado, Margaret Beach, Eileen Pelayo, John A. Chiorini, William F. Craft, David A. Ostrov, Vijay Ramiya, Sukesh Sukumaran, Apichai Tuanyok, Blake M. Warner, Cuong Q. Nguyen

**Affiliations:** Department of Infectious Diseases and Immunology, College of Veterinary Medicine, University of Florida, Gainesville, Florida, USA; Salivary Disorder Unit, National Institute of Dental and Craniofacial Research, NIH, Bethesda, Maryland; Laboratory of Pathology, Center for Cancer Research, National Cancer Institute, NIH, Bethesda, Maryland; AAV Biology Section, National Institute of Dental and Craniofacial Research, NIH, Bethesda, Maryland, USA; Department of Comparative, Diagnostic, and Population Medicine, College of Veterinary Medicine, University of Florida, Gainesville, Florida, USA; Department of Pathology, Immunology & Laboratory Medicine, College of Medicine, University of Florida, Gainesville, Florida, USA; LifeSouth Community Blood Centers, Gainesville Fl; Valley Children’s Hospital, Madera, California; Department of Oral Biology, College of Dentistry; Center of Orphaned Autoimmune Diseases, University of Florida, Gainesville, Florida, USA

**Author notes:** Authors contributed equally to the study (Yiran Shen, Alexandria Voigt). Co-correspondence authors. Address correspondence: Cuong Q. Nguyen, PhD, Department of Infectious Diseases and Immunology, PO Box 110880, College of Veterinary Medicine, University of Florida, Gainesville, Florida 32611-0880. USA, Blake M. Warner, DDS, PhD, MPH, Salivary Disorders Unit, Building 10 Room 1A01, 10 Center Drive, National Institutes of Health, Bethesda, MD 20895.

**Keywords:** Sjögren’s Disease (SjD), Severe Acute Respiratory Syndrome Coronavirus 2 (SARS-CoV-2), Coronavirus disease 2019 (COVID-19), Autoimmune disease, Autoantibodies

## Abstract

**Objectives:** Sjögren’s Disease (SjD) is a chronic and systemic autoimmune disease characterized by lymphocytic infiltration and the development of dry eyes and dry mouth resulting from the secretory dysfunction of the exocrine glands. SARS-CoV-2 may trigger the development or progression of autoimmune diseases, as evidenced by increased autoantibodies in patients and the presentation of cardinal symptoms of SjD. The objective of the study was to determine whether SARS-CoV-2 induces the signature clinical symptoms of SjD.

**Methods:** The ACE2-transgenic mice were infected with SARS-CoV-2. SJD profiling was conducted. COVID-19 patients’ sera were examined for autoantibodies. Clinical evaluations of convalescent COVID-19 subjects, including minor salivary gland (MSG) biopsies, were collected. Lastly, monoclonal antibodies generated from single B cells of patients were interrogated for ACE2/spike inhibition and nuclear antigens.

**Results:** Mice infected with the virus showed a decreased saliva flow rate, elevated antinuclear antibodies (ANAs) with anti-SSB/La, and lymphocyte infiltration in the lacrimal and salivary glands. Sera of COVID-19 patients showed an increase in ANA, anti-SSA/Ro52, and anti-SSB/La. The male patients showed elevated levels of anti-SSA/Ro52 compared to female patients, and female patients had more diverse ANA patterns. Minor salivary gland biopsies of convalescent COVID-19 subjects showed focal lymphocytic infiltrates in four of six subjects, and 2 of 6 subjects had focus scores >2. Lastly, we found monoclonal antibodies produced in recovered patients can both block ACE2/spike interaction and recognize nuclear antigens.

**Conclusion:** Overall, our study shows a direct association between SARS-CoV-2 and SjD. Hallmark features of SjD salivary glands were histologically indistinguishable from convalescent COVID-19 subjects. The results potentially implicate that SARS-CoV-2 could be an environmental trigger for SjD.

**Key Messages:** What is already known about this subject?

- SAR-CoV-2 has a tropism for the salivary glands. However, whether the virus can induce clinical phenotypes of Sjögren’s disease is unknown.

What does this study add?

- Mice infected with SAR-CoV-2 showed loss of secretory function, elevated autoantibodies, and lymphocyte infiltration in glands.
- COVID-19 patients showed an increase in autoantibodies. Monoclonal antibodies produced in recovered patients can block ACE2/spike interaction and recognize nuclear antigens.
- Minor salivary gland biopsies of some convalescent subjects showed focal lymphocytic infiltrates with focus scores.

How might this impact on clinical practice or future developments?

- Our data provide strong evidence for the role of SARS-CoV-2 in inducing Sjögren’s disease-like phenotypes.
- Our work has implications for how patients will be diagnosed and treated effectively.

## Introduction

Sjögren’s Disease (SjD) is an autoimmune disease that is generally categorized by sicca symptoms in the mouth and eyes, the presence of autoantibodies, and lymphocytic infiltration into the salivary gland[1,2]. It is estimated that approximately 4 million Americans are affected, making SjD the second most common autoimmune disease after rheumatoid arthritis[3–5]. SjD has the most skewed sex distribution known, with a 9:1 ratio of women to men[6]. SjD is most closely associated with symptoms of dryness, particularly of the mouth and eyes; however, a wide variety of extraglandular manifestations have been reported involving virtually any organ or tissue[4,7]. The extraglandular manifestations of SjD have been subdivided into visceral (gastrointestinal tract, lungs, heart, central and peripheral nervous system) and non-visceral (muscles, joints, skin) involvement, indicating the wide variety of tissues that may be involved in the disease. While both men and women at any age can be affected by SjD, it is most commonly diagnosed in women in the fourth or fifth decade of life[7,8]. The pathological framework of SjD pathogenesis remains elusive, however studies have suggested the primary drivers are genetic susceptibility, hormonal factors, and environmental triggers.

In December 2019, a novel coronavirus, severe acute respiratory syndrome-coronavirus-2 (SARS-CoV-2), emerged in Wuhan, Hubei Province, China, initiating a breakout of atypical acute respiratory disease, termed coronavirus disease 2019 (COVID-19). SARS-CoV-2 is a *betacoronavirus* in the family of *Coronaviridae*; the virus contains four structural proteins: S (spike), E (envelope), M (membrane), and N (nucleocapsid), sixteen non-structural proteins (nsp1−16) and eleven accessory proteins, which support viral essential physiological function and evasion from the host immune system[9]. As of May 1^st^, 2022, approximately one million U.S. residents have died from COVID-19[10], with more than 80 million total cases. Recent studies have identified the association between SARS-CoV-2 infection and autoimmune response. A recent literature review[11] (n= 1176 articles and 90 case reports) revealed that the primary rheumatic diseases associated with COVID-19 patients were vasculitis, arthritis, idiopathic inflammatory myopathies, and systemic lupus erythematosus. Several studies have found an association between antinuclear antibodies (ANAs) (35.6%) and COVID-19 infection, where the leading reactive antigens include SSA/Ro (25%), rheumatoid factor (19%), lupus anticoagulant (11%), and type I interferons (IFN-I) (10%)[12–14]. In 6 independent case studies, COVID-19 patients were diagnosed with systemic sclerosis[15], adult-onset Still’s disease[16], sarcoidosis, and systemic lupus erythematosus (SLE) with 4/6 patients acutely manifesting during COVID-19. An elevated level of anti-SSA/Ro52 in COVID-19 patients was linked to pneumonia severity and poor prognosis[17]. The underlying mechanism for the production of autoantibodies in COVID-19 patients is unknown, however, it poses a significant challenge for post-COVID-19 symptoms or post-acute sequelae of SARS-CoV-2 (PASC).

There are reports and cases of COVID-19 patients experiencing ocular and oral symptoms. Keratoconjunctivitis was observed in a few patients during a specific phase of the disease[18]. One study has shown that xerostomia was observed in 29% of the patient cohort[19] while another showed an increase of 30% in reporting xerostomia during hospitalization[20]. While these early studies had small sample sizes, the results appeared to indicate an association between COVID-19 and oral and ocular manifestations, primary symptoms of SjD. Increased rates of xerostomia in this patient cohort may be explained by tropism of SARS-CoV-2 for the salivary glands, resulting in host immune response and immune-mediated injury[21]. Furthermore, growing evidence of autoantibody production in COVID-19 patients raises a critical question as to whether SARS-CoV-2 infection is a risk factor for primary SjD. Therefore, the goal of this study was to determine the autoimmune response triggered by SARS-CoV-2 infection. The results indicate that infection with SARS-CoV-2 recapitulated an SjD-like phenotype in transgenic mice. Additionally, we show by using human sera paired by sex, age, and race that not only do they contain general autoantibodies but those associated with SjD, namely anti-SSA/Ro52 and anti-SSB/La.

## Methods

### Human samples

SARS-CoV-2 positive and healthy control (HC) sera were obtained from the CTSI Biorepository at University of Florida in compliance with IRBs 202001475 and 2020000781. The presence of SARS-CoV-2 was confirmed by RT-PCR for admittance into the CTSI Biorepository Bank. Peripheral blood mononuclear cells (PBMC) from five post-convalescent COVID-19 donors were obtained from LifeSouth Community Blood Centers (Gainesville Fl). The healthy volunteer donors had recovered from COVID-19 and were positive for SARS-CoV2 antibodies at the time of blood donation. The donors had no prior clinically diagnosed autoimmune diseases. Handling of the samples was performed in a certified BSL2+ with Institutional Biosafety Committee approved protocols.

#### NIDCR Subjects and Protocols

Subjects were consented to National Institutes of Health (NIH) Central Institutional Review Board (IRB)-approved protocols (15-D-0051: *Characterization of Salivary Gland Disorders* [PI-Warner]; 20-D-0094: *Transmissibility and Viral Load of SARS-CoV-2 in Oral Secretions* [PI-Warner]) and evaluated at either the NIH SARS-CoV-2 Field Testing Facility (20-D-0094) or the NIH Clinical Center. NIH IRB Protocol: 15-D-0051 (NCT02327884), is a cross-sectional screening protocol to evaluate subjects with a variety of disorders affecting the salivary complex and also, healthy subjects (i.e., healthy volunteers [HV]). All enrolled subjects are evaluated comprehensively including: oral, sialometric, ophthalmologic, and rheumatologic evaluations; salivary gland ultrasonography, bloodwork including rheumatologic investigations, and minor salivary gland (MSG) biopsies. NIH IRB Protocol: 20-D-0094 (NCT04348240) was a short-term longitudinal study aimed at examining the potential transmissibility and viral load of SARS-CoV-2 in saliva when compared with nasal and nasopharyngeal secretions, and for testing the effectiveness of masks to reduce speaking-related transmission[21]. The general results of this study are reported in Huang, et al., (2021)[21]. After identifying SARS-CoV-2 in saliva, the protocol was amended to allow MSG biopsy in acute and convalescent COVID-19 subjects[21].

Research and clinical records post-initiation of the global COVID-19 pandemic were reviewed systematically by a rheumatology Physicians Assistant (MB). Subjects were included in the histopathological analysis if they had recovered from COVID-19, had convalescent MSG biopsies, and were enrolled on NIH IRB Protocols: 15-D-0051 or 20-D-0094. Subjects were excluded if they were evaluated as a patient for the workup for SjD or non-SjD sicca symptoms. Comprehensive investigations as described above were completed on subjects enrolled on our 15-D-0051 protocol, but due to constraints of the NIH SARS-CoV-2 Field Testing Facility, these parameters were not able to be collected on all 20-D-0094 subjects. Clinical laboratory studies at NIH include standard bloodwork, assays for antinuclear antibodies (ANA), antibodies to extractable nuclear antigens (e.g., anti-SSA/SSB autoantibodies), and antibodies to pathogens to assess vaccination and exposure history purposes (e.g., anti-spike, anti-nucleocapsid). In one subject (subject 2), serial MSG biopsies were collected; the first was taken 5 days after the first COVID-19 symptoms (reported previously as COV49[21]) and the second was taken 6 months later[21].

MSG biopsies were interpreted by a board-certified anatomic pathologist (DEK) for diagnostic purposes and the histopathology was systematically reviewed by a board-certified oral and maxillofacial pathologist (BMW) as previously described (PMID: 30996010). Salivary gland inflammation and fibrosis were graded according to Greenspan et al. [PMID: 4589360] and Tarpley et al. [PMID: 4586901]. For MSG with Greenspan grade 3 or 4 sialadenitis, a focus score was calculated according to Daniels et al. [PMID: 1055974]. Hematoxylin and eosin [H&E], CD20, CD3, CD4, CD8) was conducted by the Anatomic Pathology Laboratory of the National Cancer Institute. Slides were scanned at ×40 with a NanoZoomer S360 slide scanner (Hammamatsu Photonics, Hammamatsu-city, Japan), and digital photomicrographs at ×5 resolution were captured using NDP.view2 software (Hammamatsu Photonics).

## Results

### SARS-CoV-2 triggered the decrease in the salivary secretory function

SjD patients experience xerostomia, primarily as a result of the diminished secretory function of the salivary glands. In the spontaneous animal models of SjD, the secretory dysfunction occurs between 15-20 weeks of age. Here, we sought to determine if SARS-CoV-2 can compromise saliva secretion by the glands. The homozygous K18-hACE2 mice were intranasally inoculated with 860 PFU of SARS-CoV-2 WA1/2020 inoculum drop-by-drop into both nostrils until fully inhaled. Saliva were collected on day 21 prior to euthanization. As shown, the infected mice showed a significant loss of salivary flow rates compared to the uninfected mice (infected: 6.64 ± 1.075 vs uninfected: 13.12 ± 0.532 ul/gr) (**Figure 1A**). The loss of saliva was equivalent to approximately a 50% reduction in function (**Figure 1B**). The infected mice showed a decrease in body weight when compared to the uninfected mice; however, the decrease was not statistically significant (**Figure 1C**). The results suggest that SARS-CoV-2 infection has a negative effect on the secretory function of the salivary glands.

**Figure 1:**
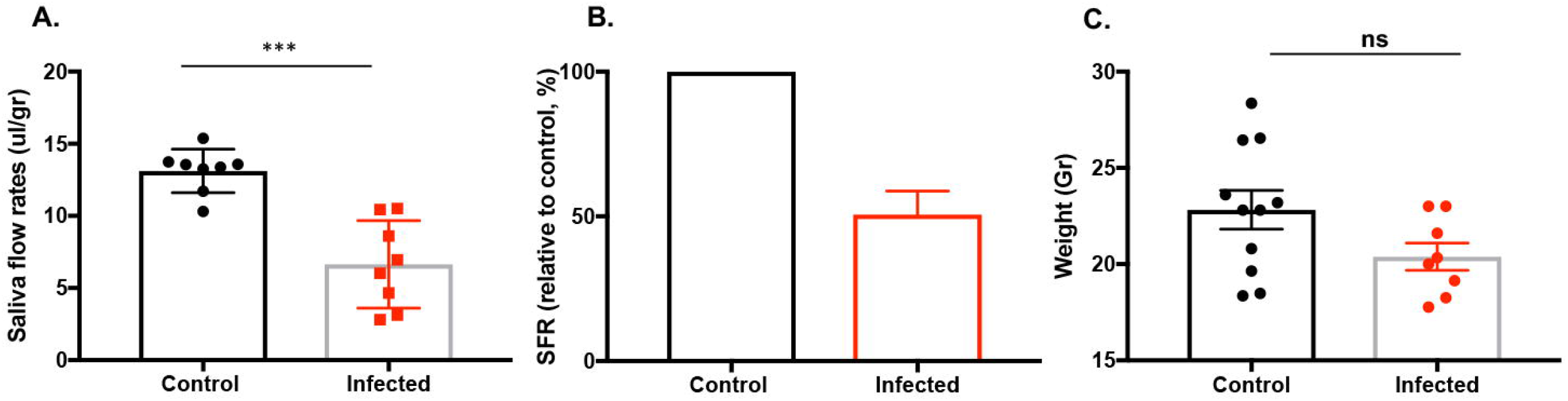
Decrease in saliva secretion by salivary glands by SARS-CoV-2. **A**. Saliva flows were collected as described in the materials and methods section. The data shown represent the saliva flow rate (ul/gram). The mice were randomly selected for saliva collection at the endpoint (control/uninfected n=8 and infected n=8). To minimize the exposure of working in BSL-3 mouse colony, the smaller number of mice was chosen for saliva collection. **B**. The mean percentage loss of saliva flow rates (SFR) in comparison to the control mice, which was set at 100. **C**. The weight of the mice in grams (control n=11, infected n=8) Data were presented as mean ± SEM. One-tailed Mann-Whitney t-tests were performed where *** p< 0.001, ns: not significant.

### SARS-CoV-2 induced the production of autoantibodies

Seropositivity for ANA and anti-SSA/Ro is one of the major classifying criteria for SJD. Here, we sought to determine if SARS-CoV-2 infection was able to induce autoantibody production. As presented in **Figure 2A**, 70% of infected mice were positive and 30% were negative for ANA using HEp2 cell staining. Whereas, in the uninfected mice, 70% were negative, and 30% were positive for ANA. Furthermore, we examined the SjD-specific autoantibodies. As indicated in **Figure 2B**, anti-SSB/La levels were highly elevated in the infected group in comparison to the control group. There was a slight, though statistically insignificant, increase in anti-SSA/Ro52 levels in the infected group. There was no change in anti-SSA/Ro60 levels between the two groups. The results suggest that SARS-CoV-2 infection in mice promotes the development of ANA and autoantibodies signature to SjD.

**Figure 2:**
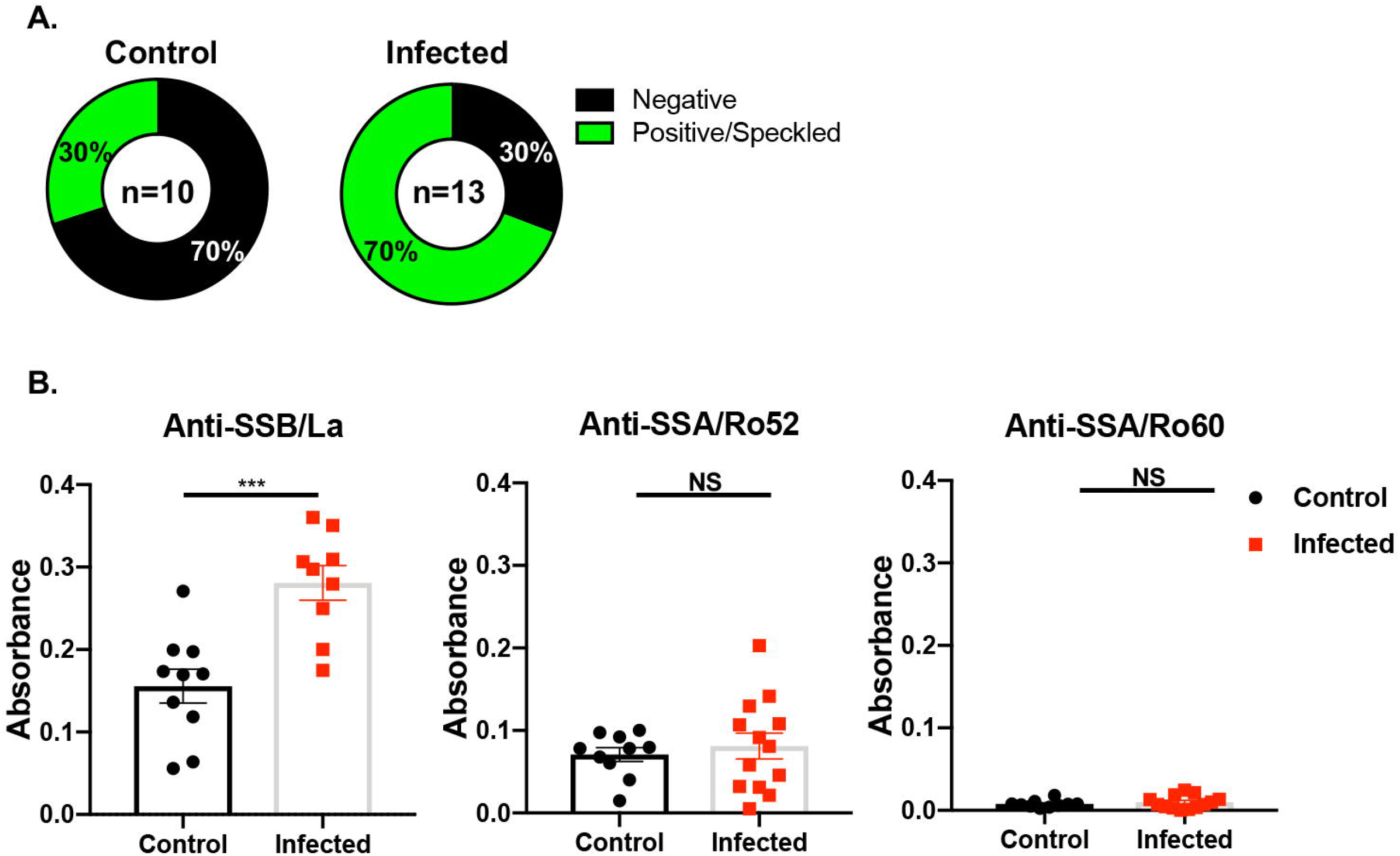
Autoantibody profile of mouse sera. **A**) ANA profile was determined using HEp2 cells. A Chi-squared test was performed on the control (n=10, 5 females, 5 males) and SARS-CoV-2 infected samples (n=13, 6 males, 7 females), with a value of 32, p <0.00001. Sera were diluted as described and positive signals were detected at 1:40-1:320 titers. **B**) Anti-SSB/La, anti-SSA/Ro52, and anti-SSA/Ro60 were determined using ELISA. Welch’s t-test was performed to determine the significance of these results, where ***p= 0.0003, ns: not significant.

### SARS-CoV-2 caused inflammation in the lacrimal and salivary glands of mice

The principal targeted tissues for SjD are the lacrimal and salivary glands. The inflammatory lesions are composed of a multitude of immune cell types, notably B cells, T cells, and macrophages. The lacrimal glands of infected mice had multifocal apoptosis/necrosis of low to moderate numbers of acinar epithelial cells characterized by cells with condensed, hypereosinophilic cytoplasm and pyknotic nuclei with karyorrhexis. The apoptosis/necrosis resulted in variable collapse and loss of acini. The interlobular duct epithelium was unaffected. The salivary glands of infected mice showed a lymphoid nodule in the interstitium, a sign of lymphocyte infiltration which was not present in control mice (**Figure 3A)**. Elevated number of apoptotic cells were found in both glands of infected mice (**Figure 3B**). Macrophages were more consistent in the salivary glands of infected mice with a smaller frequency of T or B cells. Whereas, macrophages in the lacrimal glands of infected mice were elevated drastically and consistently in the infected mice in comparison to the salivary gland. Similarly, B and T cells were detected at higher frequencies in the lacrimal than in the salivary glands (**Figure 3B**). Lymphocytic foci were detected in both the lacrimal and salivary glands of the infected mice, but not those of the control mice. Only a single infected mouse developed a focus score (FS) in the salivary glands, so deviation from the control group is insignificant (χ^2^=.27, p= 0.10247). However, FS were detected in the lacrimal glands of 5 infected mice (χ^2^=13, p= 0.00031) (**Table S1**). Additionally, lymphocytic infiltration of B and/or T cells which does not qualify as a focus was examined. This is an indicator of localized inflammation in the salivary (χ^2^=11, p= 0.00091) and lacrimal glands (χ^2^=24, p= 0.00001). Overall, SARS-CoV-2 induced inflammation with multifocal apoptosis/necrosis with more severity in the lacrimal than salivary glands.

**Figure 3:**
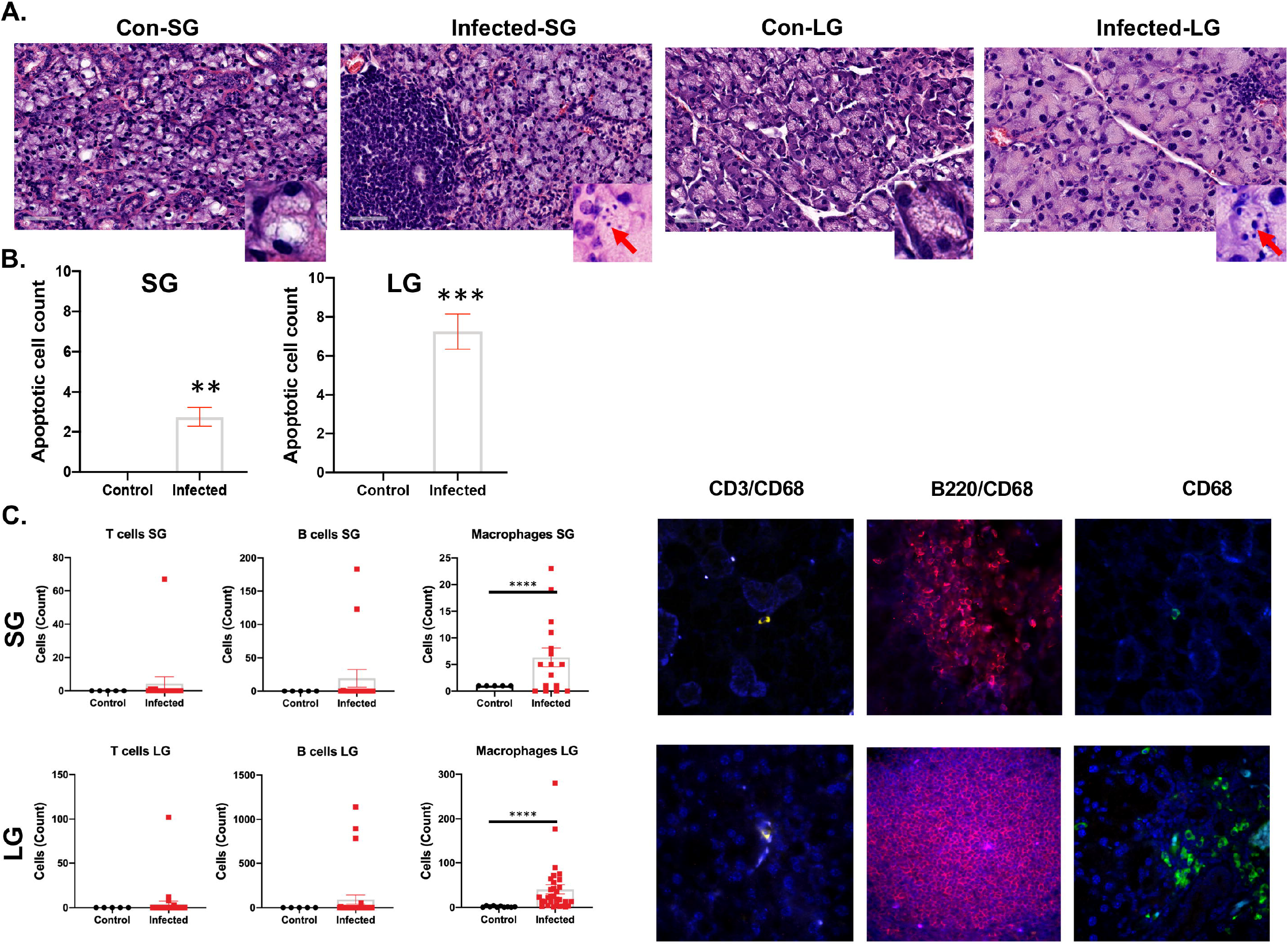
Histological examination of lymphocytes in the salivary and lacrimal glands. **A**) H&E staining of the salivary and lacrimal glands of the control and SARS-CoV-2 infected mice. Inlets with red arrows showed the apoptotic/necrotic acinar cell bodies (control, n=5 females: infected, n=26, 13 males, 13 females). **B**) Elevated number of apoptotic cells in the salivary and lacrimal glands of the infected mice. **C**) Increase in the frequency of macrophage in salivary and lacrimal glands of the infected mice. Enumeration of immune cells using immunofluorescent staining. Identification of infiltrating cells in the salivary glands and lacrimal glands, where immunofluorescent staining of CD3^+^ T cells, B220^+^ B cells and CD68^+^macrophages are displayed in yellow, red, and green, respectively, with blue DAPI nuclei staining. One-tailed Mann-Whitney t-tests were performed where ** p< 0.01, *** p< 0.001 **** p< 0.0001. Con: Control, SG: salivary glands, LG: lacrimal glands.

### COVID-19 is associated with higher autoantibody levels in a sex-specific manner

As described above, mice infected with SARS-CoV-2 developed ANA and elevated anti-SSB/La. Here, we sought to determine if these findings were also observed in human patients. As presented in **Figure 4A**, the COVID-19 patients exhibited higher frequencies of positive ANA at different sera titers compared to healthy controls. Noteably, 60% patients showed positive ANA with none for healthy controls at 1:160 titer. 30% patients still exhibited positive ANA at 1:320 titer. Further analysis of the staining patterns revealed that among the positive ANA for patients, 40% were homogeneous, 15% were speckled, and 5% were centromeric (**Figure 4B**). To further determine if the COVID-19 patients presented with SjD signature autoantibodies, patient sera were examined for reactivity against SSA/Ro52, SSA/Ro60, and SSB/La. As presented in **Figure 4C**, anti-SSA/Ro52 and anti-SSB/La were significantly elevated in COVID-19 patients in comparison to healthy controls. Anti-SSB/Ro60 levels remained similar between the two groups.

**Figure 4:**
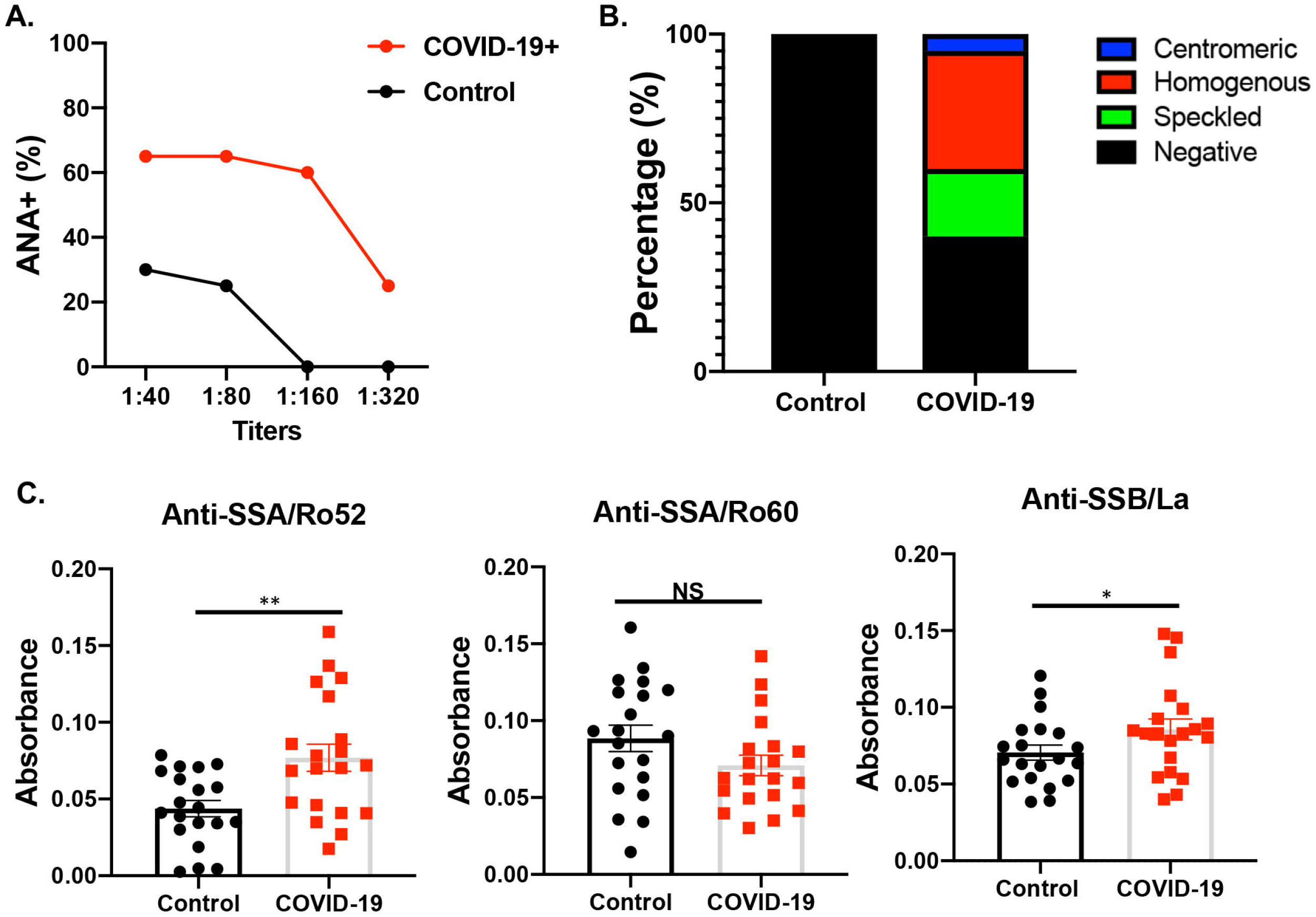
Autoantibody induction in COVID-19 human sera. **A**) ANA profile was determined using HEp2 slides at various sera titers. **B**) A breakdown by specific ANA staining pattern is presented. **C**) Anti-SSB/La, anti-SSA/Ro52, and anti-SSA/Ro60 were determined using ELISA Welch’s t-test was performed to determine the significance of these results, where **p= 0.0015 and *p= 0.0415.

SjD has a strong predilection for females; therefore, we sought to determine whether COVID-19 patients exhibited an element of sexual dimorphism in the autoantibody response. Interestingly, when examining the ANA staining it was discovered that the female COVID-19 patients had a significantly higher percentage of positive ANA at various titers compared to either the male COVID-19 patients or either sex of control patients (**Figure S1A**). Additionally, the female COVID-19 patients were shown to present a more diverse ANA pattern, with 30% speckled, 40% homogenous, and 10% centromeric at 1:160 titer, whereas the male patient showed 10% speckled and 30% homogenous pattern at the same titer. The female patient still exhibited 20% speckled with males showed 10% speckled at 1:320 titer and homogenous pattern remained the same for both sexes. positive staining for all other groups only contained a homogenous pattern (**Figure S1B**). To further determined if male and female COVID-19 patients exhibited different levels of SjD signature autoantibodies, we analyzed the samples based on sex. As presented in **Figure S1C**, female and male COVID-19 patients showed significantly higher levels of anti-SSA/Ro52 in comparison to their respective counterparts. Interestingly, male COVID-19 patients showed elevated levels of anti-SSA/Ro52 above female COVID-19 patients (p=0.0029). There was no statistically significant difference between male and female COVID-19 patients with anti-SSA/Ro60 or anti-SSB/La. The results indicated that female patients manifested more diverse patterns of ANA, however male patients exhibited higher levels of anti-SSA/Ro52 than female patients.

### Monoclonal antibodies produced by COVID-19 patients are reactive against nuclear antigens

It is remarkable to observe the cross-reactivity of COVID-19 patients’ sera against self-antigens as demonstrated. To further evaluate the B cell response of COVID-19 patients, we produced and selected nine monoclonal antibodies (mAbs) from convalescent COVID-19 patients by isolating CD20^+^ memory B cells reactive against both the RBD and S1 of SARS-CoV-2, and examined their response to self-antigens. As presented in **Figure S2A**, the mAbs exhibited various degrees of inhibition against SARS-CoV-2 RBD, in which mAbs A10 and B5 showed the highest inhibitory activity at different dilutions. To determine if they react against nuclear antigens, we tested them against HEp2 cells. As described in **Figure S2B**, seven of the nine S1/RBD-reactive mAbs produced a strong homogenous staining pattern with 100% at 1:40 and 1:80 titers and lowered to 90% at 1:160 and 1:320 titers. Overall, the results demonstrate that mAbs against the virus produced in recovered COVID-19 patients are cross-reactive and capable of recognizing nuclear antigens.

### Convalescent COVID-19 subjects demonstrate inflammation of the salivary glands and clinical signs and symptoms of Sjögren’s Disease

Six generally-healthy, relatively young (Range: 19-42y; Mean: 31y) subjects who had recovered from COVID-19, had convalescent MSG biopsies were identified for the study. These subjects were free from evidence of pre-existing autoimmune disease or major medical conditions. Subjects 1-3 were enrolled on NIH IRB Protocol: 20-D-0094 and had convalescent MSG biopsies 6-13 months after recovery from COVID-19 (Table 1). In addition, Subject 2 also received an initial biopsy during acute COVID-19 (Table 1, Figure 5). These subjects recovered from COVID-19 without continued post-acute COVID-19 symptoms as primary clinical concerns. Subjects 4-6 were enrolled on an NIH IRB Protocol: 15-D-0051 as healthy volunteers (HV) and did not present with clinical complaints of Sjogren’s Disease or post-acute COVID-19 syndrome (Table 1). Their COVID-19 status was determined from subject interviews and serological studies. In subjects with known COVID-19, their clinical course was generally mild; three of the subjects reported lung involvement with shortness of breath without hospitalization and a single subject reported significant gastrointestinal involvement (‘mild-to-moderate COVID-19’). A single subject, Subject 5 was unaware of their post-COVID-19 status and was considered ‘asymptomatic’. Evidence of infection included clinical reports of infection in five of six subjects, clinical nasopharyngeal swab PCR for SARS-CoV-2 N1 and N2 genes in four subjects (S1-3,6); anti-nucleocapsid antibodies were positive in all six subjects (*data not shown*). No subjects were postitive for anti-nuclear antibodies (ANA) or anti-SSA/Ro antibodies. A single subject was low-titre positive for anti-SSB/La antibodies. Three of the six subjects reported dry mouth during acute COVID-19 which was sustained temporarily after recovery (up to 3 weeks); a single subject had objective evidence of dry mouth (Subject 2) during acute COVID-19. Interestingly, this subject did not produce saliva from the submandibular glands for ∼3 of the 4 weeks of weekly follow up after infection. Dry eye assessments could not be completed in the NIH COVID-19 Testing Facility for three subjects, two of the three subjects who presented through the NIH Dental Clinic had objective evidence of dry eye disease (**Table 1**) but did not have clinical complaints of dry eyes.

**Table 1.**
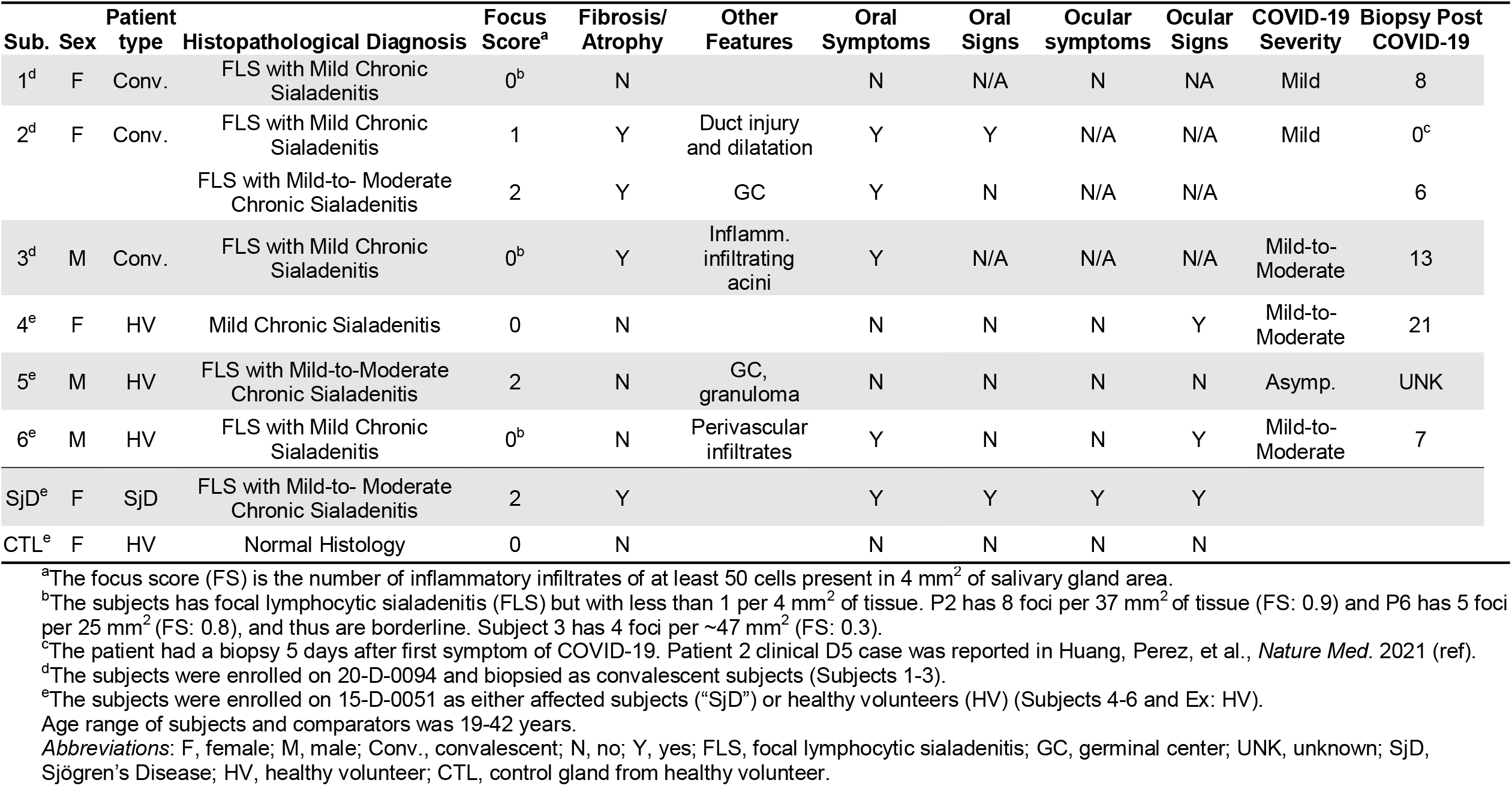
Clinical and histopathological features of convalescent COVID-19 subjects and comparators.

**Figures 5:**
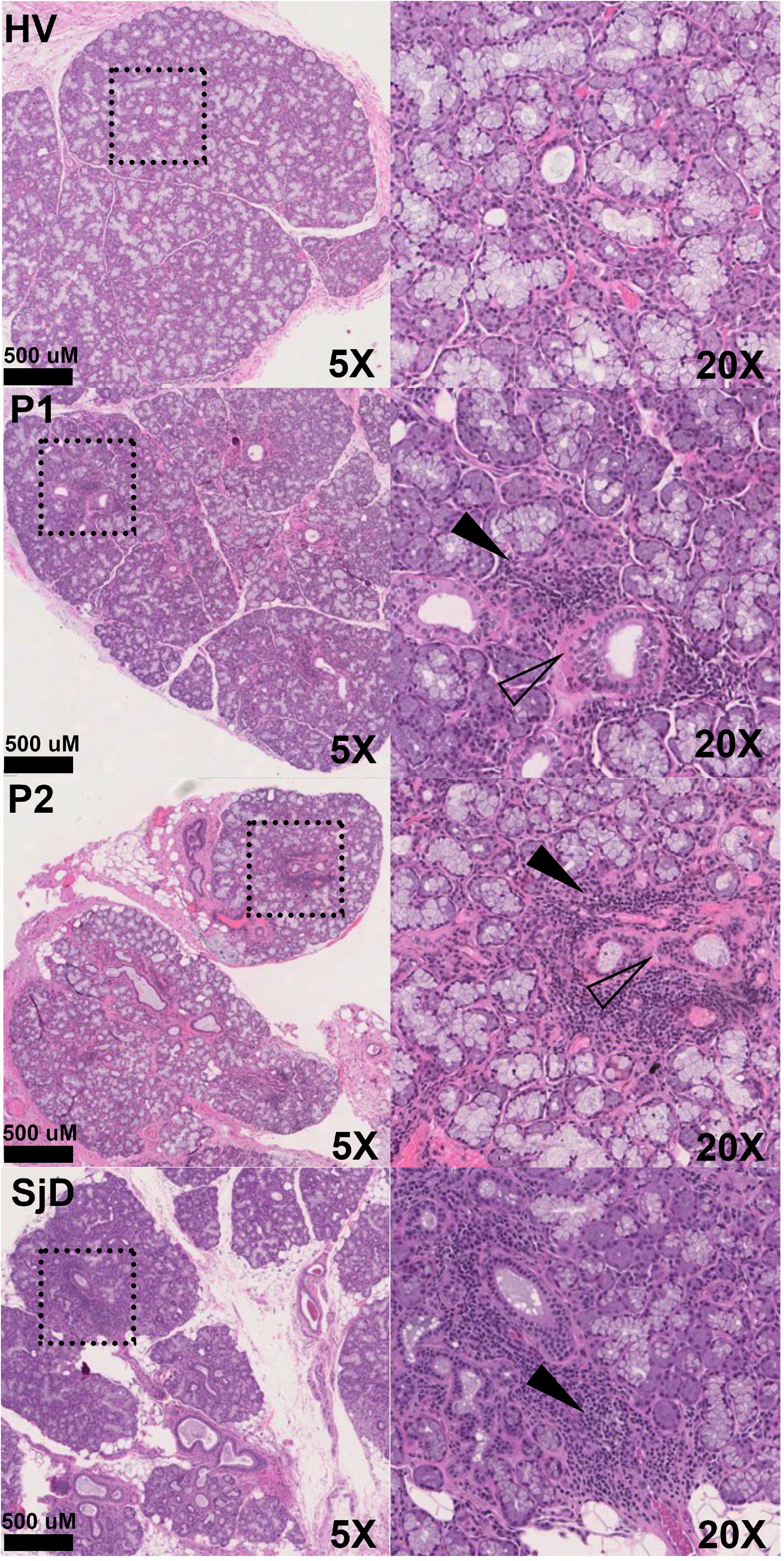
Representative minor salivary glands H&E photomicrographs of health (HV), Sjogren’s Disease (SjD), and two representative subjects recovered from COVID-19. Convalescent glands exhibit a range of inflammation severity ranging from normal to mild-to-moderate sialadenitis with focal lymphocytic sialadenitis reminiscent of inflammation found in SjD. The histopathological findings from three patients (P2 & P5) exhibit inflammation consistent with findings observed in SjD salivary glands (e.g., focal lymphocytic sialadenitis with focus scores >1.0). However, P1, P3, and P6 exhibited FLS but did not reach the threshold of >1.0 focus per 4mm^2^ of tissue.

Overall, seven MSG biopsies were collected from 6 subjects - a single subject had serial biopsies. One biopsy occurred during acute COVID-19 5 days after symptom debut, and the second 6 months after recovery. Generally, biopsies exhibited mild chronic sialadenitis (**Table 1**). However, 5 of the 7 biopsies (from four of the six subjects) had multiple foci (>50 lymphocytes) of inflammation (e.g., focal lymphocytic sialadenitis, FLS; **Table 1, Figure 5, Figure S3**). Most foci were small, although several glands exhibited multiple medium-sized and coalescing foci. Mild fibrosis and atrophy of the glands were seen in three subjects (Subject 1-3). It is noteworthy that Subject 2’s follow-up biopsy exhibited evidence of sustained immune insult as evidenced by an increased focus score (FS:1 → FS:2) and the elaboration of fibrosis and atrophy of the glands (**Figure 5**). Histopathological evidence of injury included ductal injury and mucous inspissation, immune infiltration of the acini with injury, perivascular infiltrates, and granuloma. In some subjects, the histopathological features in four of six subjects (five biopsies) are reminiscent of the range of histopathological features found in the MSG of SjD patients.

To understand the composition of the immune infiltrates, clinical immunophenotyping was performed on four biopsies from three subjects. The infiltrates generally composed of varying proportions of T cells and B cells with small foci being predominantly T cells and larger foci exhibiting a shifted balance towards B cell predominance. CD8 T cells were found both scattered throughout the gland and also in the inflammatory foci (**Figure 6, Table 2)**. These immunohistochemical studies are highly similar to the inflammatory infiltrates found characteristically in SjD. In the single subject (S2) with follow up MSG, the amount of inflammation and the shift to B cell predominance can be appreciated in the areas of FLS at 6 months.

**Figures 6:**
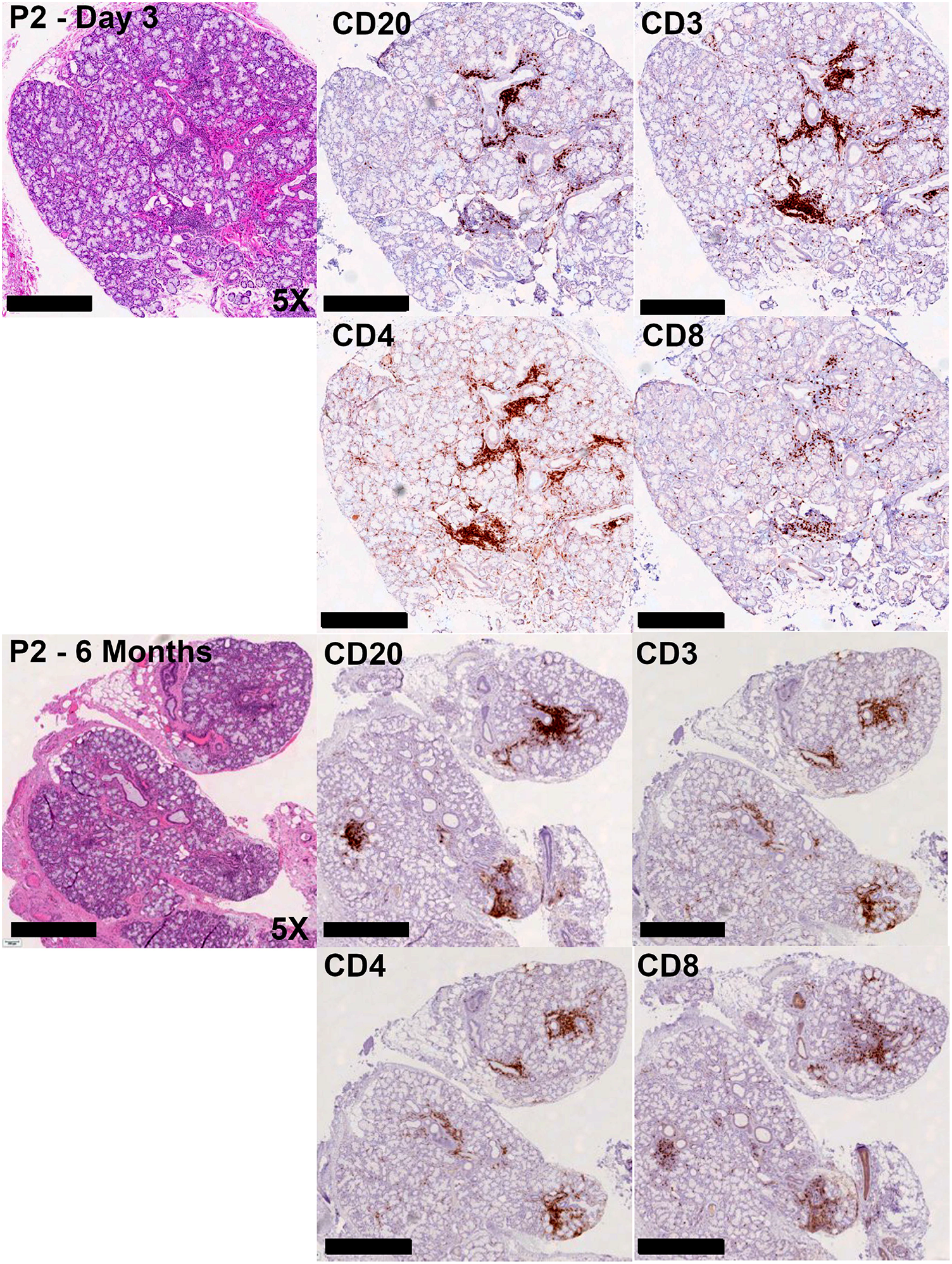
Representative immunophenotyping studies examining CD3, CD4, CD8, and CD20 on a minor salivary gland biopsy during infection (D5 post first symptom; FS: 1) and post (6 Months) COVID-19 infection (P2). Immunophenotyping demonstrates diffuse mild-to-moderate chronic sialadenitis with focal lymphocytic sialadenitis. The infiltrate is strikingly similar to infiltrates found in SjD with a predominant T cell and B cell in the ducts and acini associated with inflammation.

**Table 2.** Immunophenotyping of convalescent COVID-19 subjects salivary glands.

| Subject | CD20 | CD3 | CD4 | CD8 | CD4:CD8 |
| --- | --- | --- | --- | --- | --- |
| 1 | + | ++ | ++ | + | CD4>CD8 |
| 2-1 | + | ++ | ++ | + | CD4>CD8 |
| 2-2 | ++ | ++ | ++ | ++ | CD4>CD8 |
| 3 | + | + | + | + | CD4>CD8 |

## Discussion

Increasing evidence has supported the associations between viral/bacterial infections and autoimmune diseases. An early study demonstrated that murine cytomegalovirus induced an SjD-like disease in C57Bl/6-lpr/lpr mice with sialadenitis, severe salivary gland inflammation, and production of anti-SSA/Ro and anti-SSB/La[28]. Recent studies suggested that the virus has a tropism for the SG, including SARS-CoV-2[21,29]. Here, we sought to determine if SARS-CoV-2 infection could also trigger SjD-like phenotypes in a murine model. The results indicate that SARS-CoV-2 infection recapitulates several signature disease phenotypes, specifically, diminished salivary flow rates, salivary and lacrimal gland inflammatory lesions, and elevated autoantibodies. Similar findings were also observed in COVID-19 patients, in which, significantly elevated levels of anti-SSA/Ro52 and anti-SSB/La were seen. Additionally, female patients manifested more diverse patterns of ANA, and male patients exhibited higher levels of anti-SSA/Ro52 than female patients. In summary, the data suggest that SARS-CoV-2 infection triggered a SjD-like disease in a murine model as well as in human patients.

SARS-CoV-2 primarily uses ACE2 as a receptor[30,31], which is broadly expressed by endothelial and epithelial cells, including those of the aerodigestive tract and the salivary glands[21,32–34]. It has now been shown that salivary glands can robustly support infection and replication of SARS-CoV-2, and that saliva is potentially infectious and transmissible[21]. Intra-individual spread of SARS-CoV-2 initiates from the epithelial cells of the upper respiratory tract (e.g., acinar and ductal cells of the salivary glands) by active replication and egress of offspring viruses subsequently infecting ACE2-expressing cells in downstream organs, including the heart, kidneys, gastrointestinal tract, and vasculature[21,35,36]. The hACE2 transgenic model expressed high levels of hACE2 in the lacrimal glands and a lesser amount in the salivary glands. The tissue and cellular tropism of SARS-CoV-2 were not noticeably different between the two glands at the study endpoint. The more severe lacrimal gland inflammation and cell death could be attributed to the higher expression of hACE2, which allows for active viral infection and replication, however, the endpoint timeline was not able to capture these temporal infectious changes (**Figures S4 and S5**). ACE2 is expressed in squamous epithelial cells of the dorsal tongue, gingiva, and buccal tissue, and TMPRSS2 is expressed in taste bud cells and submandibular glands. SARS-CoV-2 was detected in SGs with higher levels in the minor SGs. In addition, saliva is a natural reservoir for viruses as one of the major fluids for viral detection. Therefore, it is not surprising that SARS-CoV-2 was found in the SGs and facilitated the inflammatory response.

The severity of COVID-19 is mediated by unregulated inflammation. During the later stage of the disease, immune-mediated damage leads to a progressive increase in inflammation. And patients with life-threatening pneumonia had neutralizing autoantibodies against IFN-ω and IFN-α[14]. As demonstrated, mice infected with SARS-CoV-2 developed higher levels of ANA and anti-SSB/La levels. Similarly, patients developed elevated levels of ANA, specifically anti-SSA/Ro52 and anti-SSB/La. In a study analyzing the sera and plasma from 64 COVID-19 patients, approximately 25% of patients exhibited an autoantibody response on average 12.3 days post-diagnosis and the reactivity was primarily to nuclear antigens, including RNP (n=8), SSA, SSB, dsDNA, chromatin, or centromere[37] Chang el al. showed that autoantibodies are present in approximately 15% of healthy controls and 50% of COVID-19 patients against commonly recognized antigens in an array of autoimmune disorders, including SSA/Ro52[38]. However, Burbelo, et al., 2022 demonstrated that a considerable fraction of the autoantibody positivity in severe COVID-19 subjects may be related to receipt of intravenous immunoglobulins (IVIG)[39]. Thus, these results suggest the need for longitudinally sampled and controlled serosurveillence need to be performed. A metanalysis revealed that the development of primary rheumatic diseases associated with COVID-19 patients were vasculitis, arthritis, idiopathic inflammatory myopathies, and systemic lupus erythematosus; overall, the association between ANAs (35.6%) and COVID-19 infection was 35.6% and the reactive antigens were found at the following rates: SSA (25%), rheumatoid factor (19%), lupus anticoagulant (11%), and IFN-I (10%)[11]. Autoantibody responses in COVID-19 patients can be influenced by sex, with men exhibiting an autoantibody response after an infection defined as at least mildly symptomatic, whereas women were prone to produce this response following an asymptomatic infection; thus autoantibody profiles are highly variable between the sexes and dependent on the disease severity[40]. It is unknown how SARS-CoV-2 infection could induce a plethora of autoantibodies, specifically autoimmune-specific antibodies. One hypothesis is that tropism of SARS-CoV-2 to vulnerable cells triggers a robust immune response that damages virally-infected cells leading to the presentation of anti-viral proteins-viral particle-antibody immune complexes to antigen presenting cells in the interstitium. A study showed that heptapeptide sharing exists between SARS-CoV-2 spike glycoprotein and human proteins, an indication of the molecular mimicry mechanism[41]. However, the spike protein does not share any homology with SjD-specific autoantigens. Therefore, it remains to be determined the underlying mechanism of autoantibody response triggered by SARS-CoV-2.

We, and others, have confirmed that salivary glands are exquisitely supportive of infection and replication of SARS-CoV-2, and saliva is an ideal secretion for inter and intra-individual spread of de novo virus[21,42]. Because of the long-hypothesized connection between viral infection and the initiation of autoimmune diseases, we examined available clinical data and minor salivary gland biopsies from convalescent COVID-19 subjects with mild-to-moderate infections. While no patients satisfied strict 2016 ACR/EULAR classification criteria, focal lymphocytic sialadenitis or clinical signs and symptoms of SjD were found in most of the available subjects[43]. In select patients, the histopathological features of inflammation in the salivary glands are indistinguishable from SjD and in the proper clinical context would be supportive of the diagnosis.

The most prevalent and persistent oral symptoms associated with COVID-19 include taste dysfunction, in addition to that, dry mouth (xerostomia) is often overlooked in COVID-19 patients and was identified as another highly prevalent (43%) oral manifestation of COVID-19[44]. A review of 12 studies, including patients diagnosed with SARS-CoV-2 infection from different countries with reported oral symptoms associated COVID-19 infection showed that xerostomia occurs in the early stages of COVID-19 with a prevalence ranging from 20% to 61.9%, and can persist for at least 8 months after recovery[45]. The percentage is higher in patients with mild symptoms, as a study in Israel showed 61.9% of 97 confirmed non-hospitalized patients reported xerostomia[46]. This is consistent with our data showing a markedly diminished salivary secretion after SARS-CoV-2 infection in mice. The precise etiology of gland dysfunction requires further investigation. As demonstrated, the influx of inflammatory cells in the glands, concomitantly with the rapid increase of acinar cell apoptosis/necrosis, may contribute to the diminished gland function. We did not measure tear secretion, mainly to avoid further physical stress on the mice as a result of the drug side effect and handling.

In summary, our study underpins the pathogenic role of SARS-CoV-2 in SjD. SARS-CoV-2 induced gland inflammation leading to the loss of saliva in mice. It triggered the production of SjD-specific autoantibodies in mice and human patients. This study raises the prospect of managing SjD in long COVID-19. Further studies are needed to examine the pathoetiology of SARS-CoV-2 in SjD.

## Data Availability

Data are available upon reasonable request

## Data availability statement

Data are available upon reasonable request

## Ethics statement

### Patient consent for publication

Not required

### Ethics approval

Subjects were consented to National Institutes of Health (NIH) Central Institutional Review Board (IRB)-approved protocols (15-D-0051: *Characterization of Salivary Gland Disorders* [PI-Warner]; 20-D-0094: *Transmissibility and Viral Load of SARS-CoV-2 in Oral Secretions* [PI-Warner]). Convalescent samples were collected under the University of Florida approved protocol (IRB202000781). Participants gave informed consent to participate in the study before taking part.

## Acknowledgments

This research was funded by the National Institutes of Health (NIH), National Institute of Dental and Craniofacial Research (NIDCR), Division of Extramural Research (DE028544, PI-Nguyen; DE028544-02S1, PI-Nguyen), and the NIH/NIDCR Division of Intramural Research (Z01-DE000704, PI-Warner; Z01-DE00695, PI-Chiorini). We give our special thanks to the members of the NIDCR Sjögren’s Disease Clinical Research Team for their coordination of patients and collection of research data and tissues.

