## Supplemental data for "Evidence of a Sjögren’s disease-like phenotype following COVID-19"

*Authors contributed equally to the study

[#]Co-correspondence authors.

Address correspondence:

Cuong Q. Nguyen, PhD

Blake M. Warner, DDS, PhD, MPH

Salivary Disorders Unit

Building 10 Room 1A01

10 Center Drive

National Institutes of Health

Bethesda, MD 20895

### METHODS

#### Animal models and $LD_{50}$ determination:

The homozygous K18-hACE2 mice were bred at the University of Florida (Breeding Project ID #3276). Mating pairs of hemizygous K18-hACE2 mice were purchased from Jackson Laboratory (034860–B6.Cg-Tg(K18-ACE2)2Prlmn/J). F1 mice were anesthetized for ear punching. The ear punch samples were collected for DNA extraction and subjected to zygosity analysis. The F1 zygosity (homozygosity) was confirmed via an SNP assay (The Jackson Laboratory's Protocol 38275: Sanger sequencing assay – Chr2_rs13476660-SEQ) by Transnetyx Inc. This assay detects an SNP between C57BL6/J and SJL/J (homozygous mutant), located approximately 4.7 kb from the transgene integration site. Once the homozygosity of the F1 mice was confirmed, mice between 8-10 weeks old were moved into micro-isolator cages, 5 mice per cage, under pathogen-free conditions for a challenge in the ABSL-3 laboratory. After being acclimated in the ABSL3 laboratory for three days, the mice were anesthetized with 87.5 mg/kg of ketamine and 12.5 mg/kg of xylazine of body weight by intraperitoneal (ip) injection. Once fully anesthetized, each group of mice was intranasally inoculated with 20 µL of SARS-CoV-2 inoculum drop-by-drop into both nostrils until fully inhaled. The mice received 860 PFU of SARS-CoV-2 WA1/2020 (BEI Resources, NR-52281) (n=26). The naïve control mice (n=10) were inoculated with 20 µL of Dulbecco's Modified Eagle Medium (DMEM) supplemented with 2% fetal bovine serum (FBS). The mice were observed twice daily until the end of the study for 21 days. The mice were euthanized when moribund. The $LD_{50}$ value was calculated using Probit analysis.

#### Making viral inoculum

One 150 $cm^2$ flasks (T-150) containing 75-80% confluent monolayer of Vero E6 cells grown in Minimum Essential Medium (MEM) Eagle supplemented with 10% heat-inactivated FBS was brought from the BSL-2 laboratory into a BSL3 laboratory two days before the experiment and incubated at 37°C with 5% $CO_2$ in an incubator. A frozen stock of 2 mL of SARS-CoV-2

WA1/2020 culture (BEI Resources, NR-52281; Centers for Disease Control and Prevention) was thawed at 37ºC. After aspirating the culture medium from the T-150 flask, the thawed viral culture was inoculated onto the monolayer of the Vero E6 cells. The flask was incubated for 1 hour. The viral inoculum was aspirated off. A freshly prepared 25 mL of MEM Eagle supplemented with 2% heat-inactivated FBS was added to the flask. The flask was incubated at the same growth condition as above. The monolayer was checked daily for signs of cytopathic effect. After 4 days of the infection, the viruses were harvested from the culture medium. The culture medium containing cells and the viruses was collected in a 50 mL conical tube. The tube was centrifuged at 4000 x g for 5 minutes to pellet the cells. The supernatant was collected and aliquoted into 2 mL vials before being stored in -80ºC freezer until used. A solid double overlay plaque assay was used to quantify the viral particles as previously described[22].

**Immunofluorescent staining for B Cells, T Cells, and macrophages**

Paraffin-embedded tissues were deparaffinized by pressure cooking in Trilogy (Cell Marque, Rocklin, CA) according to the manufacturer's instructions. Staining for B Cells, T Cells, and macrophages were performed as previously described[23]. Stained samples were visualized using 400x magnification of Nikon Ti-E fluorescent microscope with an exposure of 200 milliseconds.

**Histological examination of salivary and lacrimal glands**

Whole mouse heads were fixed in 10% phosphate-buffered formalin for at least 48 hours, then salivary and lacrimal glands were excised, embedded in paraffin, sectioned at a thickness of 5 μm, and stained as described previously[23]. Human MSG were fixed immediately after biopsy in 10% neutral-buffered formalin for 24 hours, embedded in paraffin, sectioned at a thickness of 5 μm, and stained with hematoxylin and eosin. Histopathological assessment was performed and reviewed by a board-certified surgical pathologist (DK) and an oral and maxillofacial pathologist

(BMW). When available, immunohistochemistry was performed to characterize immune infiltrates in the MSG using CD3 (T cells), CD20 (B cells), CD4 (CD4 T cells), and CD8 (CD8 T cells); the intensity and pattern of the infiltrates was interpreted.

**Detection of antinuclear antibodies**

Antinuclear antibody detection in the sera of mice and humans and mAbs was performed per the manufacturer's instructions (Immuno Concepts, Sacramento, CA). Briefly, sera were diluted 1:40-1:320 in PBS and incubated on HEP-2 ANA slides for 30 minutes. For mAbs, undiluted samples were added where concentrations were all below 1.5 mg/ml. Goat anti-mouse IgG AF488 or goat anti-human IgG AF488 were added as secondary antibodies (Invitrogen, Waltham, MA). Slides were sealed with Vectashield DAPI medium (Vector Laboratories, Burlingame, CA) and a glass coverslip was mounted. ANA staining pattern was observed at 400x with a Nikon Ti-E fluorescent microscope with an exposure of 200 ms (Nikon, Tokyo, Japan).

**Measurement of saliva flow**

To measure stimulated flow rates of saliva, individual mice were weighed and given an intraperitoneal (ip) injection of 100 µl of a mixture containing isoproterenol (0.2 mg/1 ml of PBS) (Sigma-Aldrich, St. Louis, MO) and pilocarpine (0.05 mg/1 ml of PBS) (Sigma-Aldrich, St. Louis, MO). Saliva was collected for 10 min from the oral cavity of individual mice using a micropipette starting one minute after injection of the secretagogue. The volume of each saliva sample was measured and calculated in relation to the mouse's weight to produce saliva flow rate (SFR).

**Immunohistochemical evaluation of SARS-CoV-2**

Slides were deparaffinized, then antigens were retrieved using Antigen Unmasking Solution (Vector Laboratories, Burlingame, CA) per the manufacturer's instructions. Slides were blocked with goat sera in Avidin block (Vector Laboratories, Burlingame, CA), followed by mouse anti-SARS-CoV-2 Nucleocapsid protein (Bioss, Woburn, MA) at 1:100 dilution with Biotin block (Vector Laboratories, Burlingame, CA) and incubated for 15 mins. After incubation with the secondary antibody (biotinylated goat anti-mouse IgG, Vector Laboratories, Burlingame, CA), ABC reagent (Vector Laboratories, Burlingame, CA) was added followed by DAB staining (Vector Laboratories, Burlingame, CA). Counterstaining with hematoxylin was performed. Slides were scanned through Leica microscope at 400X. The results were analyzed on Aperio ImageScope viewing software (v12.4.3.5008, Leica).

**Monoclonal antibody generation from convalescent COVID-19 patients**

RBD (ACROBiosystems, Newark, DE) and S1 proteins (Novus Biologicals, Littleton, CO) were labeled with Dylight 488 and 650 (ThermoFisher Scientific, Waltham, MA), respectively per the manufacturer's instructions. Convalescent patient PBMCs were stimulated as previously described[24] and cells were stained with DAPI, anti-CD20 PE (Biolegend, San Diego, CA), and 1:1,000 dilutions of the aforementioned SARS-CoV-2 proteins. Live, CD20+ B cells reactive against both RBD and S1 were sorted with the Sony SH800 Sorter (Minato City, Tokyo, Japan) into lysis buffer (Takara Bio Inc, Kusatsu, Shiga, Japan). Additionally, some heavy chains were recovered in a similar fashion by using SCAN[24] in which the capture slides were coated with anti-IgG and anti-Ig; captured antibodies were measured for reactivity to these proteins. Live, B cells reactive against RBD were manually picked with a micromanipulator. The variable (V) heavy (H) ($V_HDJ_H$) and light (L) ($V_LJ_L$) chains of immunoglobulins (Igs) genes of individual cells were amplified using primer sets adapted from Rodda et al[25] and Tiller et al[26] for In-Fusion Cloning Kit (Takara Bio Inc, Kusatsu, Shiga, Japan). To express Igs, VH and VL were cloned into expression vectors pFUSEss-CHIg-hG1, pFUSE2ss-CLIg-hL2 and pFUSE2ss-CLIg-hK, respectively (Invivogen, San Diego, CA). The ligation products were transformed into competent Stellar™ Competent Cells and selected by zeocin and blasticidin resistance. Cloned plasmids were sequenced through GENEWIZ (Azenta, Chelmsford, MA) and aligned by NCBI IgBLAST to ensure sequences of chains were intact. To produce monoclonal antibodies (mAbs), FreeStyle™ CHO-S cells were transfected with corresponding H/L expression vector DNA through FectoCHO® Expression System (PolyPlus, Vectura, France) as instructed by the manufacturer. On day 7 post-transfection, the whole cell culture was centrifuged and purified through EconoFit UNOsphere SUPrA on NGC™ Chromatography Systems according to the manufacturer (Bio-Rad, Hercules, CA). Protein concentration was quantified by Bradford protein assay and the integrity of the Ig fractions with joined H/L chains was determined by Mini-PROTEAN TGX Stain-Free Precast Gels (Bio-Rad, Hercules, CA) by sodium dodecyl sulfate–polyacrylamide gel electrophoresis (SDS/PAGE).

**Validation of neutralizing abilities of mAbs through competitive ELISA**

The anti-SARS-CoV-2 neutralizing antibody titer serological assay kit (AcroBiosystems, Newark, DE) was used to measure the neutralizing capacity of mAbs according to the manufacturer. The samples were diluted to 2ug/mL, 1ug/mL and 0.5ug/ml. The samples were added to wells containing 0.3μg/mL of RBD-conjugated with HRP that binds to the immobilized human ACE2 protein pre-coated on 96-well plates. The final absorbance was detected at 450 nm after adding substrate solution and subsequently stop solution. The results of the detected signal were normalized to the percentage of inhibition by comparing to the positive control, and a percentage of inhibition of 20% was considered to be inhibitory. CR3022 (NR-52481, BEI) was used as a neutralizing antibody control.

**Detection of SjD-specific autoantibodies in sera**

Detection of human and mouse anti-SSA/Ro52, anti-SSA/Ro60, and anti-SSB/La was performed as previously described[27]. Absorbance values of 450/630 nm were determined with a Tecan Infinite M200 Pro-spectrophotometric plate reader (TECAN, Mannedorf, Switzerland).

**Statistical analysis**

Statistical analyses were performed using Prism 8 software (GraphPad, La Jolla, CA). Where indicated, 2-way ANOVA, Welch's t-tests, or Mann-Whitney t-tests were performed. In all cases, $p$ values $< 0.05$ were considered significant. For the ANA staining, a Chi-squared test was performed.

### RESULTS

#### Table S1: Focal scores in salivary and lacrimal glands

| | Mice (n) | Focal scores | | | | | | Lymphocytic infiltration | | | | | |
|---|---|---|---|---|---|---|---|---|---|---|---|---|---|
| | | Salivary glands | | | Lacrimal glands | | | Salivary glands | | | Lacrimal glands | | |
| | | Positive (n) | χ2 | p-value | Positive (n) | χ2 | p-value | Positive (n) | χ2 | p-value | Positive (n) | χ2 | p-value |
| Control | 5 | 0 | 2.7 | 0.10247 | 0 | 13 | 0.00031 | 0 | 11 | 0.00091 | 0 | 24 | 0.00001 |
| Infected | 16 | 1 | | | 5 | | | 5 | | | 7 | | |

**Supplemental Table 1**: Incidence of exocrine-gland SjD symptoms in SARS-CoV-2 mice. The number of mice positive for a focal score (FS) in the salivary or lacrimal glands is provided, where an aggregate of ≥ 50 lymphocytes qualifies as a single focus. Additionally, lymphocytic infiltration of B and/or T cells which does not qualify as a focus is reported for both the salivary and lacrimal glands. A McNemar's test was performed to compare the divergence of symptomology between the control and infected groups, reported as the $\chi^2$, with p-values provided.

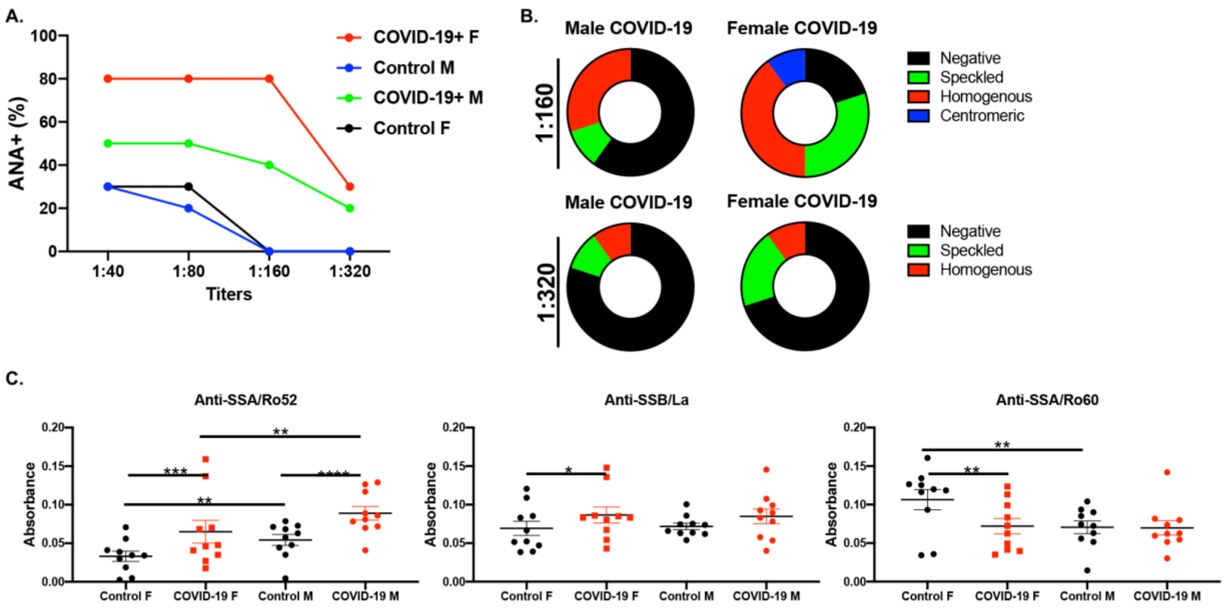


**Figure S1: Sexual dimorphic autoantibody profiles of COVID-19 patients. A**) The ANA frequencies of male and female patients and healthy controls were determined at various titers (n=10/group/sex). **B**) Staining patterns were determined at 1:160 and 1:320 titers where negative, speckled, homogenous, and centromeric staining patterns are indicated. **C**) Anti-SSB/La, anti-SSA/Ro52, and anti-SSA/Ro60 levels are presented for males and females. Two-way ANOVA was performed to determine the significance of these results, where **p<0.001, ***p= 0.0001, and ****p<0.000.

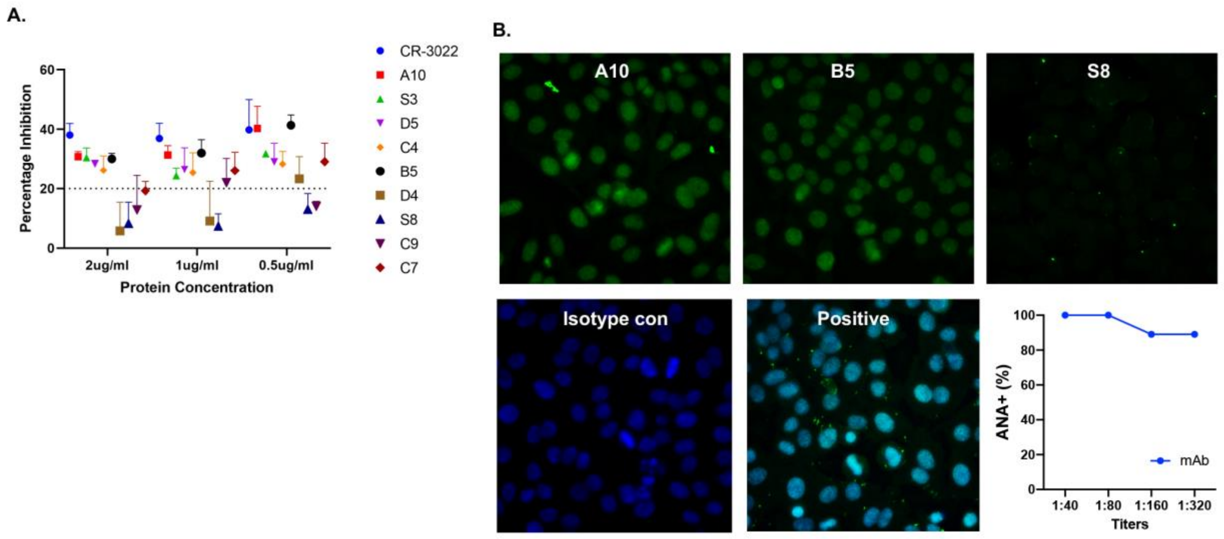


**Figure S2: Monoclonal antibodies from recovered COVID-19 patients showed reactive against self-antigens. A**) Competitive ELISA demonstrated the comparable inhibition ability of SARS-CoV-2 RBD of nine mAbs under different concentrations. CR-3022 served as positive control. Dotted line is the threshold value of the inhibition effect determined by the manufacturer. **B**) Nuclear antigen reactivity was examined using HEp2 cells. Seven of the nine S1/RBD-reactive mAbs produced a strong homogenous staining pattern at 1/40-1/320 titers. Representative images of three mAbs (A10, B5, and S8) are presented at 1/320 titer with the blue DAPI turned off to better demonstrate the homogenous pattern stained by AF488. Isotype control and positive control were shown with DAPI and AF488.

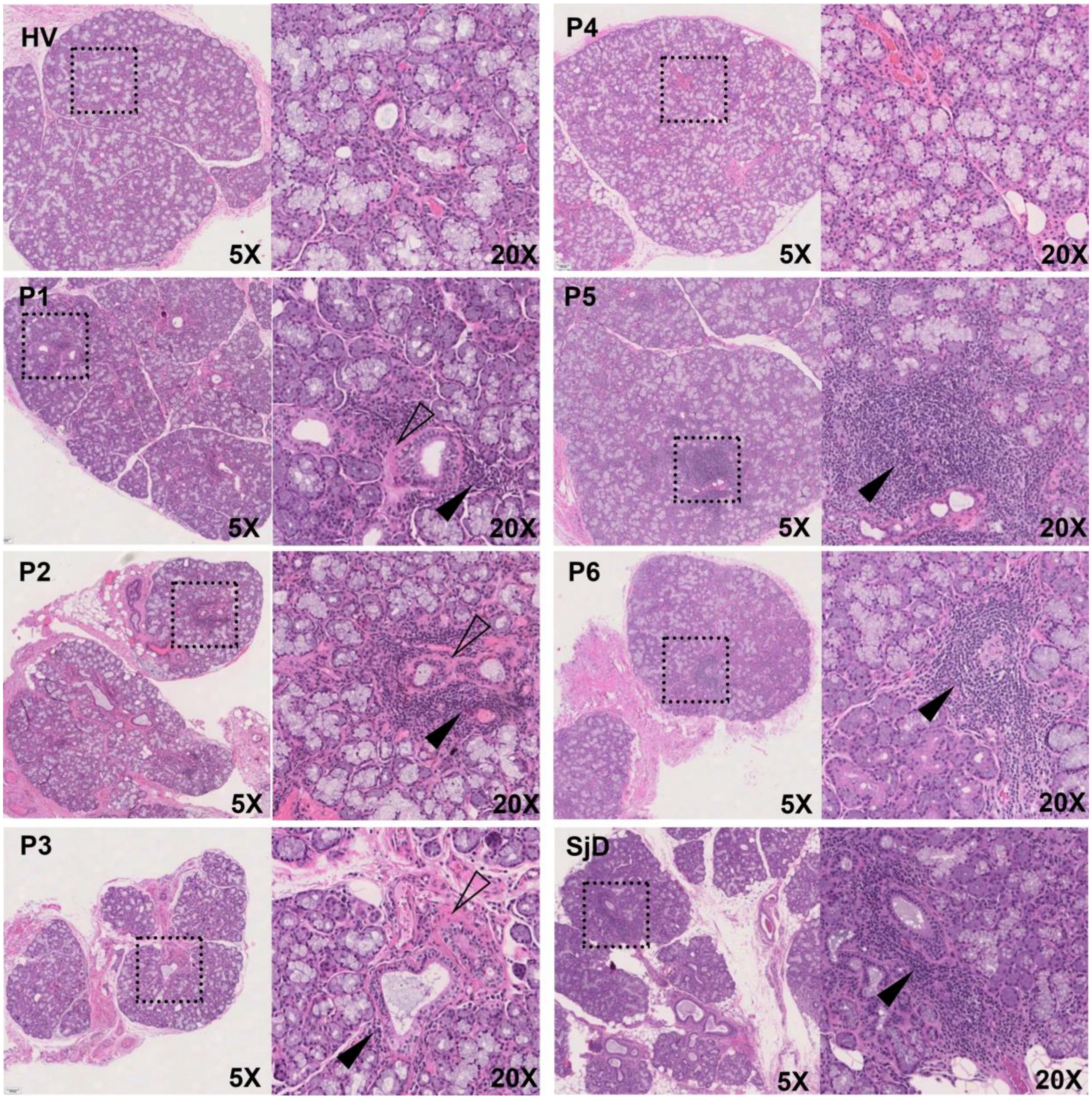


**Figure S3: Hematoxylin and eosin (H&E) staining of salivary glands obtained from 6 patients recovered from COVID-19 compared to a healthy volunteer and a Sjogren's Disease (SjD) patient.** Black arrows point to foci of inflammation and outlined arrows point to areas of fibrosis. It is noteworthy that P1, 2, 5, 6 all demonstrate FLS, although only 2/4 >1.0 foci/4mm$^2$.

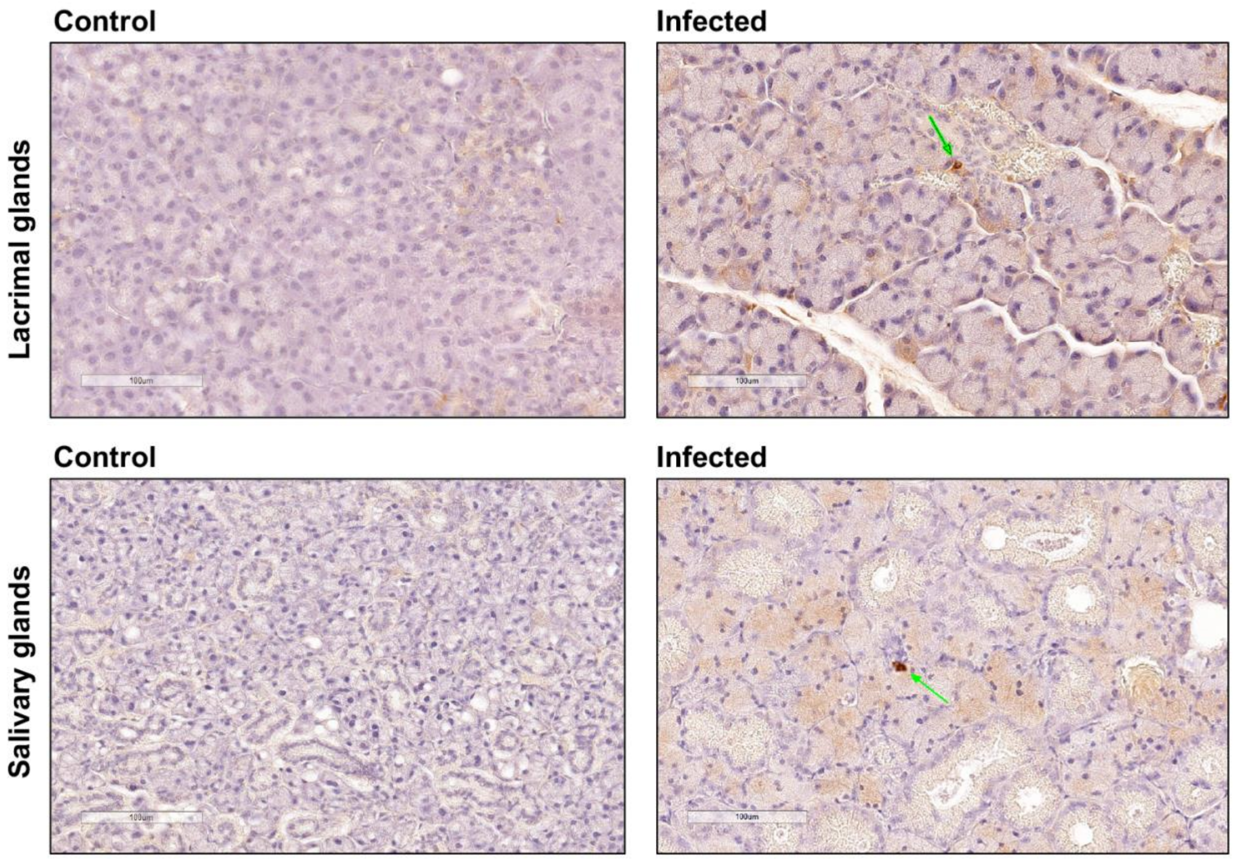


**Figure S4: SARS-CoV-2 IHC staining in lacrimal glands (LG) and salivary glands (SG) from mice infected with SARS-CoV-2**. Immunohistochemical analysis for SARS-CoV-2 nucleocapsid protein was performed with positive (arrow) in the LG and SG of infected mice. Control mice or uninfected mice showed no positivity for SARS-CoV-2 nucleocapsid. Original magnifications 200X; bar = 100 μm.

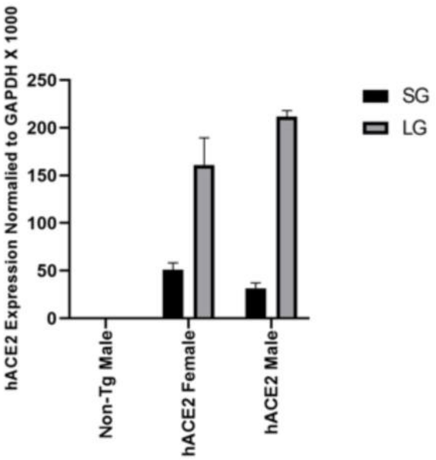


**Figure S5. hACE2 expression levels in the salivary and lacrimal glands.** Expression of human ACE2 (hACE2) mRNA in salivary glands (SG, black) and lacrimal glands (LG, grey) of control C57BL/6 mice or non-transgenic and K18-hACE2 transgenic mice was determined by quantitative RT-PCR.
